# Can PowerPoint Save Lives? Assessment of Equity and Reach of Traditional Dissemination Channels in Geriatric Trauma Education—A Mixed-Methods Study using Digital Analytics

**DOI:** 10.1101/2025.02.27.25323036

**Authors:** Ashley N. Moreno, Ariel W. Knight, Adam Nelson, Jessica Anderson, Min Ji Kwak, Linda Reinhart, Tasce Bongiovanni, Ilinca Barsan, Caroline Buck, Victoria Bandera, Sasha D. Adams, Bellal Joseph, Deborah M. Stein, Holly M. Holmes, Richard Lewis, Lacey N. LaGrone

## Abstract

**Background:** Creation of resources for evidence-based clinical guidance does not necessarily improve patient outcomes without deliberate dissemination and implementation. We sought to better understand the effectiveness of pre-recorded didactics in improving knowledge as well as the reach of various information dissemination strategies.

**Methods:** A mixed-methods study was conducted amongst all United States’ trauma care clinicians from March to August 2023. A 30-minute didactic video on anticoagulant management in geriatric traumatic brain injury (TBI), along with pre-and post-video knowledge surveys, was circulated via email, web, and social media accounts affiliated with 7 professional trauma societies as well as state and regional health departments. Digital analytics were captured, and descriptive and regression analyses conducted.

**Results:** The didactic video was viewed 1,407 times on YouTube with a mean view time of 7 minutes. A total of 311 participants responded to the survey. Most were registered nurses (RNs) from urban, non-academic institutions with 11+ years of clinical experience. Only 16.1% of respondents correctly answered all pre-video knowledge questions (n=48/298); this increased to 51.6% post-video (n=94/182). Surgeons and advanced practice providers (APPs) (r=0.148, p<0.05), male respondents (r=0.131, p<0.05), and clinicians at higher-level trauma centers (r=-0.140, p<0.05) answered more pre-video questions correctly. With regard to general knowledge acquisition practices, surgeons and APPs were more likely than RNs to report primary use of literature (r=0.134, p<0.05) and point-of-care medical information (r=0.133, p<0.05).

**Conclusion:** Viewing a recorded didactic was associated with improved knowledge of anticoagulation management in geriatric TBI. Trauma society-sponsored email is an effective means of information dissemination to urban, Level I/II trauma centers, but still fails to reach many rural and Level III/IV trauma centers. Given the low rate of correct pre-video answers and increase after viewing, further study is needed to better understand end-user needs to optimize dissemination and implementation of up-to-date clinical guidance.

**Level of Evidence:** Level IV; Survey and Web Analytics

## Background

Successful dissemination and implementation of evidence-based clinical guidance is essential for the delivery of high quality, contemporary clinical care. Although effectiveness research is abundant in the academic trauma community, the real-world facilitators of, and barriers to, information dissemination remain relatively unclear.^1,2^ Limited data suggest that some proportion of healthcare clinicians tend to utilize presentations, grand rounds-type lectures, and conferences to learn new evidence-based practices.^3^ Furthermore, inequalities exist within traditional communication channels, and issues with language-translation, paywalls, and access prevent resources from getting to frontline healthcare clinicians.^3^ As professional societal recommendations evolve, numerous additional channels are warranted for more timely dissemination and implementation of best clinical practices such that they are readily accessible to all trauma care clinicians, regardless of practice setting.

Geriatric patients (≥65 years old) represent an increasing percentage of the United States’ trauma population.^4^ Their diminished physiologic reserve, greater pre-injury comorbid disease, and more frequent polypharmacy (most notably including anticoagulants and antiplatelet agents) may exacerbate injury sequelae and contribute to increased morbidity and mortality relative to younger patients with similar injuries. Traumatic brain injury (TBI) is a particularly illustrative example, in which older age is associated with increased mortality and worse functional outcomes.^5^ While they comprise only 10% of all TBI cases, older adults account for 50% of TBI-related mortalities, some of which may be prevented with more widespread standardization of care. TBI thus denotes a prime opportunity to optimize geriatric trauma care, particularly for those with more limited healthcare resources and access to medical knowledge. However, while numerous clinical interventions have been shown to improve geriatric medical outcomes, there has been no formal assessment of dissemination strategies within the scope of geriatric trauma care.^6^

As such, we used a virtual lecture focused on anticoagulation in geriatric TBI patients to assess the effectiveness of clinical resource dissemination to trauma clinicians as well as the equity of reach of existing communication channels. In doing so, we have hypothesized that: (1) some dissemination routes will more efficiently reach trauma clinicians than others; (2) clinicians will score significantly higher on comprehension questions post-education relative to pre-education; and (3) that the clinician population reached will not mirror the intended target population of non-academic, non-Level I/II trauma center designation, non-urban clinicians. By better understanding the effectiveness of various dissemination strategies, the American Association for the Surgery of Trauma (AAST) and other professional societies can selectively utilize efficacious tactics to disseminate information to improve the national delivery of geriatric trauma care.

## Methods

This mixed-methods study was determined exempt from IRB review (Colorado Multiple Institutional Review Board, CB F490; COMIRB#: 22-0209). All United States trauma care clinicians, inclusive of nursing, advanced practice providers (APPs), therapists, and physicians practicing at level I-IV trauma centers and critical access centers (i.e., non-verified trauma centers) were eligible for inclusion. This study was informed by the Framework for Spread, which was used to organize conceptualization of audience and information interactions and linkages.^7^

YouTube and Bit.ly video views and survey data were the primary web analytics sources. The American Association for the Surgery of Trauma (AAST) Geriatric Committee created a 30- minute video entitled, “Traumatic Brain Injury and Elderly Patients on Oral Blood Thinners,” presented by Drs. Sasha Adams and Adam Nelson. The didactic video was then uploaded to YouTube in “unlisted” form, and thus could only be accessed via a shared link. A unique Bit.ly link was chosen for each virtual dissemination channel to track the number of times a specific YouTube video link was accessed (also reported as “clicks”). Bit.ly data provided participant location information (city and country of access). YouTube data yielded the overall number of video views, hours of watch time, mean view duration, and type of device being used for viewing.

The study’s cross-sectional pre-/post-video survey component conforms to the Consensus-Based Checklist for Reporting of Survey Studies (CROSS) checklist (**Supplemental Item 1**).^8^ It contains approximately 30 items that elucidate the viewer’s demographics and typical and preferred educational acquisition strategies as well as the effectiveness of content transfer through pre-and post-video knowledge questions. REDCap was used to administer the web-based pre- and post-video surveys.^9^ Survey links were provided via a QR code embedded in the video presentation and in the YouTube video description box. To ensure participant anonymity, no identifiable information was requested, and the pre- and post-video surveys were not linked. Data collection occurred from March to August 2023. A series of standardized sharing templates were created to disseminate the educational materials across various platforms (e.g., email, social media, etc.). Organization-specific communicators were encouraged to add additional context and messaging to optimally address their respective audiences. We invited participation via email from various professional trauma societies as well as state and regional health departments. Following an introductory email containing the project overview and sharing templates in March, the study team sent periodic follow-up reminders to encourage frequent dissemination until August. The American Association for the Surgery of Trauma (AAST), Society of Trauma Nurses (STN), The American College of Surgeons (ACS), American Trauma Society (ATS), Western Trauma Association (WTA), Eastern Association for the Surgery of Trauma (EAST), and Coalition for National Trauma Research (CNTR) all served as dissemination channels. In addition, states with high proportions of older adults living in rural areas were identified and publicly available contacts for their state health departments, Emergency Medical Services, and/or regional trauma systems coordinators were contacted. Additionally, presentation of the didactic video during institution-sponsored grand rounds was encouraged.

Following the close of data collection, REDCap, Bit.ly, and YouTube data were extracted and aggregated. Location data of the top six performing Bit.ly links (AAST Social Media, ATS Email, STN Email, STN Social Media, WTA Email, and State/General Regional Health Departments) were extracted for geographic analysis, and accounted for nearly 94% of all Bit.ly clicks.

Univariate, bivariate, and multivariate analyses were employed to evaluate improved geriatric trauma care knowledge among clinicians. Descriptive statistical analysis provided an understanding of variations within both the independent and dependent variables. Correlation analysis was used to determine bivariate relationships between the independent variables and knowledge growth variables.

We additionally employed multiple linear regression analysis to explore the multiple effects of independent variables on the knowledge growth variables. Seven independent variables were used: type of institution; type of clinician; years of training; location where the respondent practices; level of trauma center in which the respondent practices; Gender; and Race. Two knowledge growth variables were examined, knowledge growth through contact with colleagues and attending professional conferences.

Excel and IBM Statistical Package for the Social Sciences (SPSS), version 27, was used to conduct all descriptive, bivariate, and multivariate statistical analyses.

## Results

### Web Analytics

Four primary communication channels were utilized over a six-month period: email, in-person national and regional conferences, social media platforms, and organizational websites. The 30- minute didactic video was viewed 1,407 times on YouTube with a mean view time of 7 minutes. A total of 311 respondents completed the pre-video survey, and 182 respondents completed the post-video survey. Most respondents were from urban, non-academic institutions, and nearly 75% were nurses. Most were relatively experienced with over 60% having completed clinical training 11+ years ago. Surgeons (72%) and APPs (45%) were more likely to work at Level I trauma centers compared to nurses (30%) (p<0.002) (**Table 1**).

**Table 1.** Demographics of the Pre-Video and Post-Video Respondents.

| Variable | Pre-Video |  | Post-Video |  |
| --- | --- | --- | --- | --- |
|  | Number | % | Number | % |
| **Institution Type** |  |  |  |  |
| Academic | 94 | 30.2 | 56 | 30.8 |
| Non-academic | 151 | 48.6 | 80 | 44.0 |
| Academic-affiliated | 59 | 19 | 39 | 21.4 |
| Other | 7 | 2.3 | 7 | 3.8 |
| **Work Location** |  |  |  |  |
| Urban | 157 | 51.6 | 4 | 57.1 |
| Rural | 97 | 31.9 | 3 | 42.9 |
| Peri-urban | 45 | 14.8 | 0 | 0 |
| Other | 5 |  | 0 | 0 |
| **Institution Level** |  |  |  |  |
| Level I | 106 | 34.1 | 4 | 57.1 |
| Level II | 85 | 27.3 | 0 | 0 |
| Level III | 44 | 14.1 | 2 | 28.6 |
| Level IV | 52 | 16.7 | 4 | 14.3 |
| Other/NA | 24 | 7.7 |  |  |
| **Provider Type** |  |  |  |  |
| Surgeon | 26 | 8.4 | 15 | 8.2 |
| Advanced practice provider | 24 | 7.7 | 11 | 6.0 |
| Nurse | 231 | 74.5 | 139 | 75.5 |
| Other | 29 | 9.4 | 19 | 10.3 |
| **Years Since Completed Clinical Training** |  |  |  |  |
| 0-5 years | 56 | 18 | 36 | 19.7 |
| 6-10 years | 45 | 14.5 | 29 | 15.8 |
| 11-20 years | 89 | 28.6 | 47 | 25.7 |
| More than 20 years | 121 | 38.9 | 71 | 38.8 |
| **Gender** |  |  |  |  |
| Woman | 241 | 77.5 | 6 | 85.7 |
| Man | 63 | 20.3 | 1 | 14.3 |
| Non-binary/non-conforming/Prefer Not to Answer | 7 | 1.2 | 0 | 0 |
| **Race/Ethnicity** |  |  |  |  |
| White | 258 | 70.9 |  |  |
| Hispanic or Latino | 21 | 5.8 |  |  |
| Black or African American | 13 | 3.6 |  |  |
| Other* | 17 | 4.6 |  |  |
| Prefer Not to Answer | 17 | 4.7 |  |  |

\*American Indian, Alaskan Native, Asian, Native Hawaiian, or Other Pacific Islander

In analysis of the top performing Bit.ly dissemination channels (AAST Social Media, ATS Email, STN Email, STN Social Media, WTA Email, and General Regional), half (50%) of viewers were confirmed from the United States (most commonly Texas, California, and Oregon), and nearly 6% confirmed from international communities. The remaining 44% of Bit.ly accessors were from cities with fewer than five clicks/scans and thus were aggregated by Bit.ly as ‘no data’.

### Channel Reach and Devices Used

Most respondents accessed the education materials via email (85%) followed by social media (5%) and other sources (5%) (**Table 2**). Email yielded the highest average view duration (9:13 minutes) and greatest overall watch time (152.8 hours) compared to other media platforms (**Table 3**). Respondents were most likely to view educational content on either a computer (75.1%) or a mobile phone (24%) with computers yielding a longer average view duration (8:10 minutes via computer vs. 3:12 minutes via mobile phone) and greater overall watch time (143.9 hours via computer vs. 18 hours via mobile phone).

**Table 2.** Approach Which Led Respondent to View the AAST Geriatric Trauma TBI & Blood Thinners Education Video Frequency Distribution.

| **Category** | **Pre-Video** |  | **Post-Video** |  |
| --- | --- | --- | --- | --- |
|  | **Number** | **%** | **Number** | **%** |
| E-mail | 240 | 76.9 | 4 | 57.1 |
| National/Regional Conference | 5 | 1.6 | 0 | 0.0 |
| Social Media | 5 | 1.6 | 1 | 14.3 |
| Website/Banner | 26 | 8.4 | 1 | 14.3 |
| Other | 36 | 11.5 | 1 | 14.3 |
| Total | 312 | 100.0 | 7 | 100.0 |

**Table 3.** YouTube Watch Time by Organization and by Medium (N=1,407)

|  | **Views** | **Total**<br>**Watch**<br>**Time (Hrs)** | **Average**<br>**View**<br>**Duration\*** |
| --- | --- | --- | --- |
| **Organization** |  |  |  |
| AAST | 96 | 4.242 | 0:04:36 |
| ACS | 6 | 1.0084 | 0:10:05 |
| ATS | 297 | 38.568 | 0:04:12 |
| CNTR | 22 | 1.6563 | 0:04:31 |
| EAST | 17 | 1.0645 | 0:02:32 |
| General Regional | 51 | 4.4055 | 0:05:10 |
| Grand Rounds | 6 | 0.5688 | 0:05:41 |
| STN | 886 | 109.7454 | 0:04:37 |
| WTA | 26 | 3.4266 | 0:05:42 |
| **Medium** |  |  |  |
| Email | 1167 | 152.8006 | 0:09:13 |
| Other | 57 | 4.9743 | 0:05:26 |
| Podium | 7 | 0.1976 | 0:01:09 |
| Poster | 16 | 0.9562 | 0:01:57 |
| Social Media | 145 | 4.7233 | 0:01:39 |
| Website | 15 | 1.0335 | 0:04:08 |

\*The average of averages
The American Association for the Surgery of Trauma (AAST); The American College of Surgeons (ACS), American Trauma Society (ATS), Coalition for National Trauma Research (CNTR), Eastern Association for the Surgery of Trauma (EAST), Society of Trauma Nurses (STN), Western Trauma Association (WTA), "General Regional" refers to the State and General Regional Health Departments.

### Knowledge Assessment & Transfer

A total of 298 respondents completed the pre-video knowledge questions, and 182 respondents completed them post-video (**Table 4**).

**Table 4.** Pre-Video and Post-Video Frequencies of Responses to Knowledge Questions.

| **Knowledge Item** | **Category** | **Pre-Video**<br>**%** | **Post-Video**<br>**%** |
| --- | --- | --- | --- |
| *What is the impact of oral blood thinners on TBI?* |  |  |  |
|  | Incorrect Answer | 26.1 | 14.1 |
|  | Correct Answer | 73.9 | 85.9 |
|  | Total | 100.0 (310) | 100.0 (184) |
| *Role of Laboratory Coagulation Parameters in the Acute Management of Oral Blood Thinner associated TBI* |  |  |  |
|  | Incorrect Answer | 61.8 | 39.3 |
|  | Correct Answer | 38.2 | 60.7 |
|  | Total | 100.0 (301) | 100.0 (183) |
| *Which is the correct reversal agent for TBI with intracranial hemorrhage in a patient on warfarin?* |  |  |  |
|  | Incorrect Answer | 51.1 | 11.4 |
|  | Correct Answer | 48.9 | 88.6 |
|  | Total | 100.0 (305) | 100.0 (185) |
| ***Knowledge Questions Combined*** |  |  |  |
|  | 0 Correct | 9.1 | 2.2 |
|  | 1 Correct | 36.6 | 12.1 |
|  | 2 Correct | 38.3 | 34.1 |
|  | 3 Correct | 16.1 | 51.6 |
|  | Total | 100.0 (298) | 100.0 (182) |

Surgeons and APPs (r = 0.148; p <0.05) and male respondents (r = 0.131; p <0.05) had greater pre-video knowledge compared to nurses and female respondents, respectively (**Table 5**, *“Overall Knowledge”*). Clinicians at higher level trauma centers (e.g., Level I/II) had greater pre-video knowledge (r = −0.140; p <0.05) than those working at lower-level centers (e.g., Level III,IV; **Table 5**). Otherwise, no significant pre-video knowledge gaps were observed.

**Table 5.**
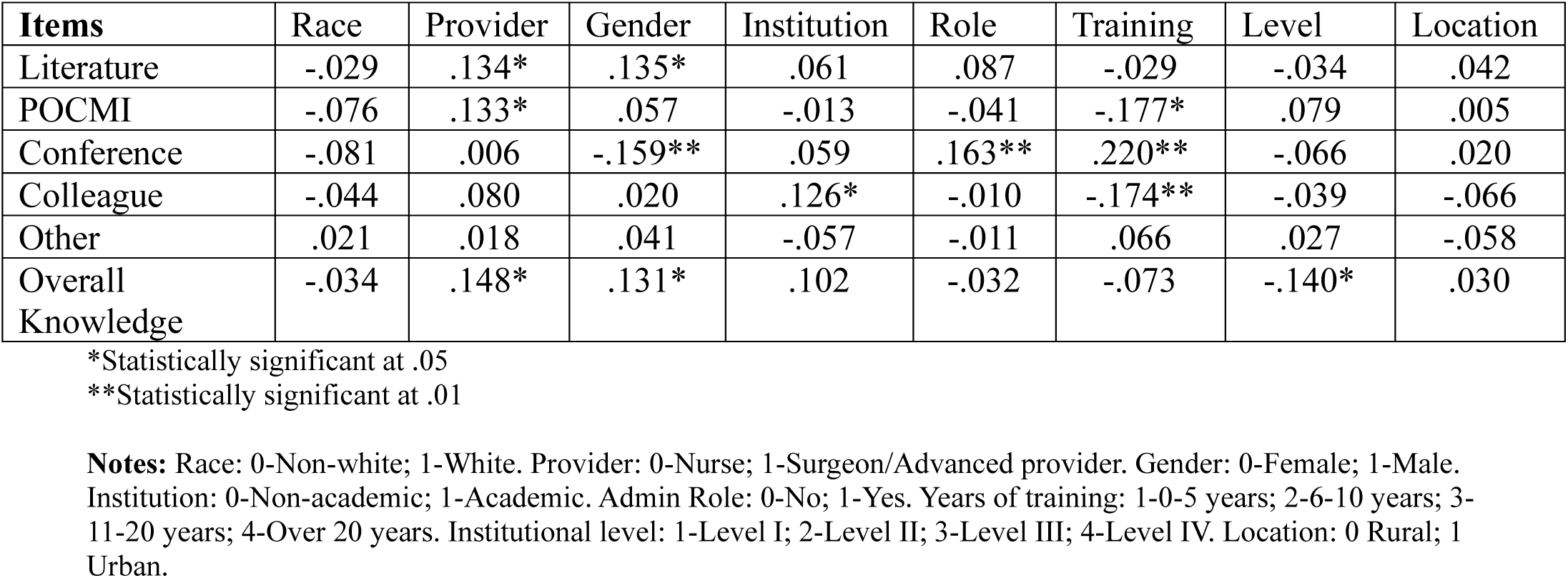
Pre-Video Correlations of Knowledge Growth and Knowledge Questions with Selected Independent Variables.

Pre-video, only 16.1% of respondents achieved 3/3 correct responses; this increased to 51.6% post-video (**Table 4**). Post-video, 34.1% of respondents achieved 2/3 correct responses. Therefore, respondents were mostly able to correctly identify the impact of oral blood thinners on TBI after completing the educational video.

### Preferred Methods of Information Access

Most respondents “usually” or “always” utilized primary literature (52%), point-of-care medical information (50%), conferences (39%), and/or a colleague to expand their knowledge (48%) (**Figure 1**). Responses did not differ by urban/rural location, academic/non-academic institutional affiliation, trauma center designation, or years of clinical experience.

**Figure 1.**
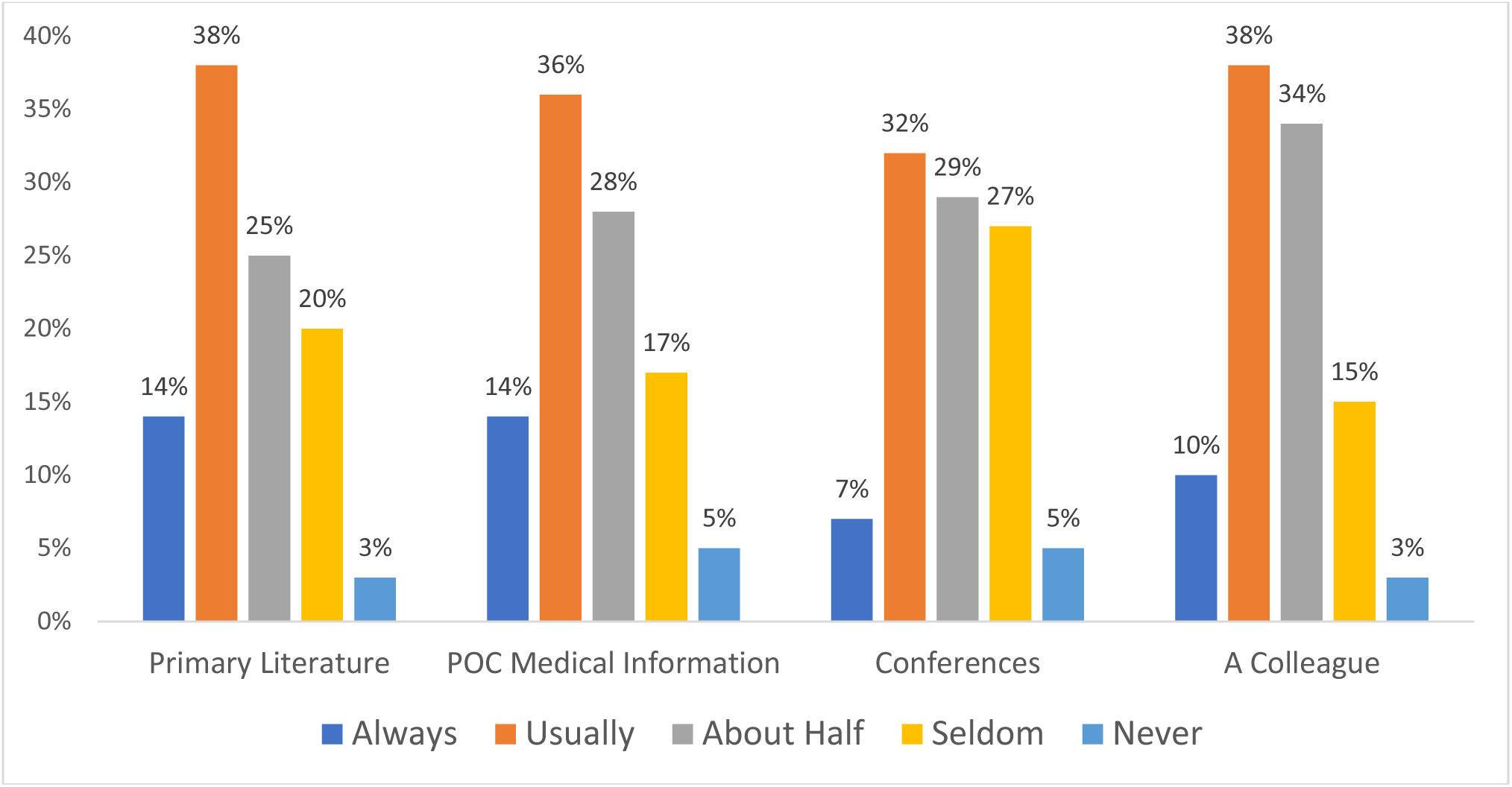
Combined Pre-Video and Post-Video Frequencies Regarding How Often the Respondent Accesses the Following Sources When Seeking Knowledge Growth.

However, surgeons and APPs were more likely than nurses to report use of primary literature (r = 0.134; p <0.05) and point-of-care medical information (r = 0.133; p <0.05) (**Table 5**). Male clinicians were more likely to report using primary literature (r = 0.135; p <0.05) while female clinicians were more likely to report acquiring information from conferences (r = −0.159; p c<0.01). Academic-affiliated respondents were more likely to confer with colleagues when they had clinical uncertainty than were non-academic clinicians (r = 0.126; p <0.05), while those in administrative roles were more likely to use conferences for knowledge acquisition than those who were not (r = 0.163; p <0.01). Clinicians with less clinical experience sought further knowledge from colleagues (r = −0.174; p <0.01) and point-of-care medical information (r = - 0.177; p <0.05) while those with more clinical experience preferentially used conferences (r = 0.220; p <0.01). Neither trauma center designation, urban/rural practice location, nor race affected the way clinicians sought additional knowledge.

### Multivariate Analysis

In multivariate analysis of pre-video responses, institution type, clinician type, years of clinical experience, practice location, trauma center designation, gender, and race were treated as independent variables. None impacted knowledge growth via access of either primary literature or point-of-care medical information.

Years of experience, gender, and race impacted the variation of reported knowledge growth through conference participation (F = 4.880; p <0.001) (**Supplemental Item 2**). Years of clinical experience had the strongest influence; as clinicians gain more independent clinical experience, they report increased knowledge growth via conference participation. Furthermore, clinicians who are women or non-white are more likely to report gaining knowledge from conference participation compared to men or white clinicians. This regression model explains nearly 13% (r^2^ = 0.126) of the variation in knowledge growth through conference participation.

Years of clinical experience influences the variation in knowledge growth through colleagues (F = 3.057; p <0.004) (**Supplemental Item 3**). As independent clinical experience increases, clinicians are less likely to seek knowledge from colleagues. The regression model explains nearly 8% (r^2^ = 0.083) of the variation in knowledge growth through colleagues.

Review of free-text items highlighted that respondents preferred to access future educational materials via either webinars (ideally with the opportunity to obtain CME credit), through a centralized website of all major trauma societal guidelines, and/or regular email newsletters.

## Discussion

Pre-recorded, virtual lectures are a practical education resource for trauma clinicians of varying levels of education, experience, academic affiliation, location, and trauma center designation. However, this study highlights numerous opportunities for improvement. The didactic video was accessed (and presumably viewed) on YouTube over 1,400 times, yet the average watch time was less than one-third of the video’s length. Paired with the low pre- and post-video survey response rates, it is likely that while many viewers’ initial interest solicited clicks, this did not yield completion of the entire module in many instances. While the module was disseminated via multiple communication channels, most clinicians accessed the video via email and viewed it on a computer. Thus, email remains a relevant educational outreach platform for this type of product, perhaps even more so than social media or society websites.

### Knowledge Assessment & Transfer

Overall, the post-video quiz yielded more correct answers compared to the pre-video quiz, likely demonstrating that clinicians, regardless of their clinical role or practice environment, are capable of learning and retaining new information to keep their knowledge and practice updated. However, since the pre- and post-video responses were not linked, these results are limited in that they cannot confirm if the completers are the same people. In addition, it is possible that these results represent a motivated minority that finished the educational video and were thus able to score well on the knowledge quiz. What these results do highlight is the need for more optimal continuing education. This may be particularly relevant to clinicians at lower-level trauma centers (e.g., Level III/IV), nurses, and those further removed from formal training.

Clinicians at Level I/II trauma centers had greater pre-video knowledge than those working at Level III/IV centers, perhaps reflecting the more comprehensive clinical resources and systems required to successfully care for more medically and surgically complex, critically injured patients at the highest-level trauma centers, as well as the fact that they more regularly care for such patients at their facilities. This may be additionally explained by disparities in access to contemporary, evidence-based protocols and the accompanying clinical resources, more regular and structured interactions with resident and/or fellow trainees who are still in formal postgraduate educational programs, and increased engagement in ongoing, clinically relevant research. Amongst clinicians, surgeons and APPs had greater pre-video knowledge than nurses. This may reflect longer, more comprehensively structured training programs for surgeons as compared to nurses. It may also simply be that trauma surgeons are expected to have a more comprehensive working knowledge of anticoagulant management in TBI than trauma nurses, despite their sharing of a mutual patient population.

Furthermore, more experienced clinicians had fewer correct pre-video answers than those with less experience. This may be explained by more recent exposure to contemporary clinical management strategies in a structured educational curriculum in a clinician’s respective specialty. As standards of care evolve, up-to-date practices may be better represented in recent graduates’ formal curricula. In analysis of patients treated by older or younger physicians, patients treated by older physicians had higher mortality.^10^

While male respondents had greater pre-video knowledge that female respondents, this observation is likely linked to clinician type rather than aptitude or another innate characteristic. In our study, surgeons were more likely to be men and nurses were more likely to be women. This reflects current gender demographics in the trauma community: women comprise less 30% of board-certified trauma/critical care surgeons as compared to nearly 80% of trauma nurses.^11,12^ Importantly, since gender was not collected for all post-video survey respondents, analysis of knowledge transfer by gender could not be conducted.

Statistically, there were no differences in pre-video knowledge based upon race, institutional academic affiliation, administration role, or rural/urban location.

### Methods of Information Access

While many trauma clinicians seek information in similar ways, interesting differences emerged. Surgeons and APPs were more likely than nurses to use primary literature and point-of-care medical information. Academic-affiliated respondents were more likely to confer with colleagues than non-academic clinicians, and those in administrative roles were more likely to utilize conferences than those who were not. Clinicians with less clinical experience sought information from colleagues and point-of-care medical information more than those with more clinical experience. Those with greater clinical experience preferentially utilized conferences. This latter preference may reflect more experienced clinicians being both presenters and consumers of conference content. More senior clinicians may also have more accrued time off and more funding to attend conferences. Furthermore, as years of training increased, the reliance on gaining knowledge from colleagues decreased. This may reflect collaborative relationships formed early on in one’s career, perhaps conferring with a more experienced colleague in a mentor/mentee dynamic. Finally, male clinicians were more likely to use primary literature while female clinicians were more likely to acquire information from conferences.

### Limitations

Our study has several limitations. Overall, the pre- and post-video survey responses were low relative to the number of people who accessed the video on YouTube. Even fewer people completed the post-video questions as compared to the pre-video questions. Therefore, our analysis may lack generalizability to the full spectrum of trauma clinicians. Additionally, while responses were deliberately not linked so as to preserve anonymity, this limited the collection of some demographic variables, the ability to track individual progress pre- and post-video, and subsequent analysis.

Based upon the available study demographics, it is likely that underrepresented populations— namely those at Level III/IV trauma centers, in rural locations, and non-white clinicians—were not sufficiently reached. For example, while over 70% of US adult trauma centers carry a Level III or lower designation, less than 30% of our respondents reported working at one, thus highlighting further outreach and educational opportunities for national trauma societies. This research had hypothesized that the clinician population reached would not mirror the intended target population of non-academic, non-Level I/II trauma center designation, non-urban clinicians. However, since this actual target population is ill-defined, we are limited in the data analyses feasible.

### Next Steps

These preliminary results of assessment of dissemination strategies to expand geriatric trauma care highlight several opportunities to formalize educational outreach and ultimately better standardize trauma care across a continuum of facilities in the United States. As many respondents favorably viewed easy access to free educational webinars, future creation of similar modules by the AAST and other national trauma organizations may allow for future CME outside of annual society meeting attendance.

While beyond the scope of this project, it is unclear if the educational module either altered clinical practices at our audience’s trauma centers or improved patient outcomes, nor were any barriers to implementation identified. These unanswered questions represent future opportunities to optimize trauma education and patient care. While this didactic module focused on geriatric TBI management, future educational content and investigations into dissemination, access, and effectiveness of such resources may be expanded to other patient populations and injury patterns to optimize the availability and utility of best practice resources by all trauma clinicians, regardless of practice setting.

## Supporting information

Supplemental Items

## Data Availability

All de-identified data produced in the present study are available upon reasonable request to the authors.

## SUPPLEMENTAL DIGITAL CONTENT

Supplemental Item 1: Consensus-Based Checklist for Reporting of Survey Studies (CROSS) checklist

Supplemental Item 2: ANOVA and Multiple Regression Summary of Pre-Video Selected Independent Variables with Knowledge Growth Through Attending Conference

Supplemental Item 3: ANOVA and Multiple Regression Summary of Pre-Video Selected Independent Variables with Knowledge Growth Through Colleagues

Supplemental Item 4: Conflict of Interest Forms

## AUTHOR CONTRIBUTIONS

Moreno: Conceptualization (supporting), Data curation (lead), formal analysis (supporting), resources (supporting), data curation (supporting), visualization (supporting), project administration (lead), writing – original draft (lead), writing – review and editing (lead)

Knight: Conceptualization (supporting), data curation (supporting), Formal analysis (supporting), writing – original draft (lead), writing – review and editing (lead)

Nelson: Conceptualization (lead), data curation (supporting), formal analysis (supporting), resources (lead), validation (supporting), visualization (supporting), writing – original draft (supporting), writing – review and editing (supporting)

Anderson: Conceptualization (lead), data curation (supporting), formal analysis (supporting), resources (lead), validation (supporting), visualization (supporting), writing – original draft (supporting), writing – review and editing (supporting)

Kwak: Conceptualization (lead), data curation (supporting), formal analysis (supporting), resources (lead), supervision (lead), validation (supporting), visualization (supporting), writing – original draft (supporting), writing – review and editing (supporting)

Reinhart: Conceptualization (lead), data curation (supporting), formal analysis (supporting), resources (lead), validation (supporting), visualization (supporting), writing – original draft (supporting), writing – review and editing (supporting)

Bongiovanni: Conceptualization (lead), data curation (supporting), formal analysis (supporting), resources (lead), validation (supporting), visualization (supporting), writing – original draft (supporting), writing – review and editing (supporting)

Barsan: Conceptualization (lead), data curation (supporting), formal analysis (supporting), resources (lead), validation (supporting), visualization (supporting), writing – original draft (supporting), writing – review and editing (supporting)

Buck: Conceptualization (lead), data curation (supporting), formal analysis (supporting), resources (lead), validation (supporting), visualization (supporting), writing – original draft (supporting), writing – review and editing (supporting)

Bandera: Conceptualization (lead), data curation (supporting), formal analysis (supporting), resources (lead), validation (supporting), visualization (supporting), writing – original draft (supporting), writing – review and editing (supporting)

Adams: Conceptualization (lead), data curation (supporting), formal analysis (supporting), resources (lead), validation (supporting), visualization (supporting), writing – original draft (supporting), writing – review and editing (supporting)

Joseph: Conceptualization (lead), data curation (supporting), formal analysis (supporting), resources (lead), supervision (lead), validation (supporting), visualization (supporting), writing – original draft (supporting), writing – review and editing (supporting)

Stein: Conceptualization (lead), data curation (supporting), formal analysis (supporting), resources (lead), supervision (lead), validation (supporting), visualization (supporting), writing – original draft (supporting), writing – review and editing (supporting)

Holmes: Conceptualization (lead), data curation (supporting), formal analysis (supporting), resources (lead), validation (supporting), visualization (supporting), writing – original draft (supporting), writing – review and editing (supporting)

Lewis: Conceptualization (supporting), data curation (supporting), Formal analysis (lead), data curation (supporting), visualization (lead), writing – original draft (supporting), writing – review and editing (supporting)

LaGrone: Conceptualization (lead), data curation (supporting), formal analysis (supporting), resources (lead), supervision (lead), validation (supporting), visualization (supporting), writing – original draft (lead), writing – review and editing (lead)

## MEETING DISCLOSURE

This work has been presented as a poster at The American Association for the Surgery of Trauma Annual Meeting in Las Vegas in September 2024.

## FUNDING ACKNOWLEDGMENTS

None

## ACKNOWLEDGEMENTS

***Wunderman Thompson***

The authors would like to thank the entire Wunderman Thompson team for their pro bono partnership in setting up the YouTube channel and all associated Bit.ly links.

***REDCap***

Study data were collected and managed using REDCap electronic data capture tools hosted at the University of Colorado. [citation below] REDCap (Research Electronic Data Capture) is a secure, web-based application designed to support data capture for research studies, providing: 1) an intuitive interface for validated data entry; 2) audit trails for tracking data manipulation and export procedures; 3) automated export procedures for seamless data downloads to common statistical packages; and 4) procedures for importing data from external sources.

***CCTSI***

This publication was supported by NIH/NCATS Colorado CTSA Grant Number UL1 TR002535. Its contents are the authors’ sole responsibility and do not necessarily represent official NIH views.

